# Machine learning algorithm to predict delirium from emergency department data

**DOI:** 10.1101/2021.02.19.21251956

**Authors:** Sangil Lee, Brianna Mueller, W. Nick Street, Ryan M. Carnahan

## Abstract

**Introduction:** Delirium is a cerebral dysfunction seen commonly in the acute care setting. Delirium is associated with increased mortality and morbidity and is frequently missed in the emergency department (ED) by clinical gestalt alone. Identifying those at risk of delirium may help prioritize screening and interventions.

**Objective:** Our objective was to identify clinically valuable predictive models for prevalent delirium within the first 24 hours of hospitalization based on the available data by assessing the performance of logistic regression and a variety of machine learning models.

**Methods:** This was a retrospective cohort study to develop and validate a predictive risk model to detect delirium using patient data obtained around an ED encounter. Data from electronic health records for patients hospitalized from the ED between January 1, 2014, and December 31, 2019, were extracted. Eligible patients were aged 65 or older, admitted to an inpatient unit from the emergency department, and had at least one DOSS assessment or CAM-ICU recorded while hospitalized. The outcome measure of this study was delirium within one day of hospitalization determined by a positive DOSS or CAM assessment. We developed the model with and without the Barthel index for activity of daily living, since this was measured after hospital admission.

**Results:** The area under the ROC curves for delirium ranged from .69 to .77 without the Barthel index. Random forest and gradient-boosted machine showed the highest AUC of .77. At the 90% sensitivity threshold, gradient-boosted machine, random forest, and logistic regression achieved a specificity of 35%. After the Barthel index was included, random forest, gradient-boosted machine, and logistic regression models demonstrated the best predictive ability with respective AUCs of .85 to .86.

**Conclusion:** This study demonstrated the use of machine learning algorithms to identify the combination of variables that are predictive of delirium within 24 hours of hospitalization from the ED.

## INTRODUCTION

Delirium is a global cerebral dysfunction seen in 8% up to 64% of patients in the acute care setting. The prevalence of delirium in the emergency department (ED) and inpatient populations is surprisingly high.^1-3^ The presence of delirium is associated with a prolonged hospital stay, a higher likelihood of skilled nursing facility placement, and a 2-to 4-fold increase in mortality.^4^ Despite the mortality rate being comparable to myocardial infarction, the fluctuating nature of symptoms, uncertainty of baseline cognitive function, and limited diagnostic modality lead to diagnostic dilemmas. By clinical gestalt alone, providers miss up to 80% of patients experiencing delirium upon presentation to the ED.^5^

Unfortunately, delirium continues to be underdiagnosed and undertreated.^5^ Although several cognitive assessment tools exist, they require additional training and dissemination. ^6-8^ Early screening and interventional options are emerging and seem promising, as reported by several recent studies.^9-11^ There is a need to identify an optimal screening strategy for delirium beyond cognitive assessment because, until we have it, delirium will likely remain an elusive diagnosis. An accurate prediction model derived from variables assessed around the time of the ED visit could be a solution to identifying patients who are at risk for delirium and may benefit the most from screening and preventive measures.

Our objective was to identify a clinically valuable predictive models for prevalent delirium within the first 24 hours of hospitalization based on the available data by assessing the performance of logistic regression and a variety of machine learning models.

## METHODS

### Study Design

This was a retrospective cohort study using data from a tertiary care medical center. Data from electronic health records for patients hospitalized from the ED between January 1, 2014, and December 31, 2019, were extracted from Institute for Clinical Translational Science Data Warehouse. The study was approved by the local institutional review board (IRB), and we adhered to the transparent reporting of a multivariable prediction model for individual prognosis or diagnosis (TRIPOD).^12^

### Participants

The study population comprised patients hospitalized from an academic Level 1 trauma center ED with approximately 60,000 visits a year. At this institution, hospitalized patients aged 65 and older were screened for delirium twice daily from the time of hospitalization until discharge. The nursing protocol was in place to assess patients with the Delirium Observation Screening Scale (DOSS), a 13-item screen observation scale for non-intubated patients. If the patient was ventilated, the Confusion Assessment Method for the ICU (CAM-ICU) was used.

Eligible patients were aged 65 or older, admitted to an inpatient unit from the ED, and had at least one DOSS assessment or CAM-ICU recorded within the first 24 hours of hospitalization. If a patient was hospitalized more than once between January 2015 and December 2019, we randomly selected an encounter defined by medical record number.

### Outcome

The outcome measure of this study was a positive delirium diagnosis within one day of hospitalization determined by a positive DOSS or CAM assessment. Twenty-four hours was chosen as the cut-off length for study inclusion to optimize the model’s ability to identify delirium cases most related to factors observed around the time of the ED visit. A positive outcome was defined as one or more positive screenings within 24 hours even if a prior assessment was negative.

### Predictors

We collected data on patient demographics, medical histories, physiological measurements, medications administered, and lab results. The following variables were used to generate and test a model: age at the time of hospitalization, sex, gender, history of stroke, dementia, severe illness defined by meeting two or more Systemic Inflammatory Response Syndrome (SIRS) criteria, transient ischemic attack (TIA), diagnosis of intracranial hemorrhage in the ED, tachypnea, and visual or hearing impairment. The physiological variables collected at the time of ED evaluation were heart rate, respiratory rate, Body Mass Index (BMI), and temperature. Medications ordered were obtained with a drug flag for opioids and benzodiazepines. We defined the anticholinergic variable as receipt of drugs classified as level 2 or 3 on an updated version of the Anticholinergic Drug Scale (Supplemental table 1 displays anticholinergics received by the sample).^13,14^ Although there is not an established Activity of Daily Living (ADL) assessment in the ED at this institution, nursing staff in inpatient units recorded a Barthel index, a measurement of the degree of assistance required by a patient determined by 10 variables describing ADL and mobility.^15^ The Barthel index was included as a continuous variable where a higher Barthel index is indicative of a higher level of independence. ADL was an important predictor for delirium in the literature but the Barthel index is not available at the time of the ED visit, so we examined the models without the Barthel index as the primary analysis and without as the secondary analysis.^16^

### Analysis

We reported summary statistics for the population by positive delirium screening. Means and Standard Deviation (SD) summarized continuous variables; frequency counts and proportions summarized categorical variables.

Missing values were imputed using KNN-imputation which has been shown to outperform other widely used imputation methods.^17^ Possible outliers were identified with the IQR Extreme Value analysis. To prevent the loss of information about variability in the study, clinical reasoning was used to determine if an outlier reflects the study population. Variance inflation factors were observed to detect multicollinearity, and continuous variables were checked for linearity by examining plots of the continuous independent variables versus the logit of the outcome.

We compared the predictive performance of five machine learning models using the Python Machine Learning library Scikit-Learn.^18^ The algorithms included Logistic Regression (LR), Decision Tree (DT), Random Forest (RF), Gradient Boosting Machine (GBM), and Gaussian Naïve Bayes (GNB), Support Vector Machine (SVM), and K Nearest Neighbor (KNN) with an intention to identify an interpretable model. Cross-validation was implemented for both hyperparameter tuning and model evaluation with AUC as the evaluation metric. This re-sampling method was selected over repeated sub-sampling to prevent any loss of information about the positive class by ensuring every observation appears in both the training and test data.

To avoid an optimistic bias that can result from using the same cross-validation procedure for both hyperparameter tuning and model evaluation, nested cross-validation was employed. In nested cross-validation, k-fold cross-validation for hyperparameter tuning is nested inside the k-fold cross-validation for model evaluation. Using tenfold nested cross-validation, the data was randomly divided into 10 equally-sized subsets. Out of the 10 sets, 9 were used to train the classifier, and the 10th was used for testing. The training set was further partitioned into 5 folds for an inner cross-validation grid search to optimize hyperparameters. This process was repeated until each of the 10 subsets had served as the test set. Similar to a regular cross-validation procedure, evaluation metrics are obtained by averaging the test set scores of the 10 runs. By conducting model selection independently in each trial of the model fitting procedure, the risk of overfitting during hyperparameter tuning is reduced. The final models were selected using a 10-fold cross-validation grid search on all available data.

### Sample Size

To examine the importance of sample size on the predictive power of a model, the performance of a gradient boosted model was tested at various sample sizes. Subsets of the data were created with stratified random sampling. At each sample size, 10-fold cross validation with 10 repeats was employed to obtain an AUC estimate and standard error. AUC estimates and confidence intervals were plotted against sample size. Increasing the sample size from 4,500 to 22,500 resulted in a .011 AUC increase. This suggests a sample size larger than the sample used in the analysis would not result in a significant increase in predictive power. However, as the sample size increased, the standard error decreased which creates more confidence in the AUC estimate. (Supplemental figure 1)

## RESULTS

### Participants

We identified a total of 60,790 unique ED encounters that resulted in hospitalization during the study period. Out of these, 24,132 of them did not have at least one DOSS or CAM-ICU recorded in their health records within the first 24 hours of a hospitalization, reducing the study population to 37,609. An additional 4,056 encounters were excluded for not having a DOSS or CAM-ICU within one day of hospitalization. After removing those with multiple encounters, the remaining 22,269 ED patients met all study criteria, and 4,955 patients had a positive delirium screening within one day of hospitalization (figure 1 and table 1).

**Table 1.**
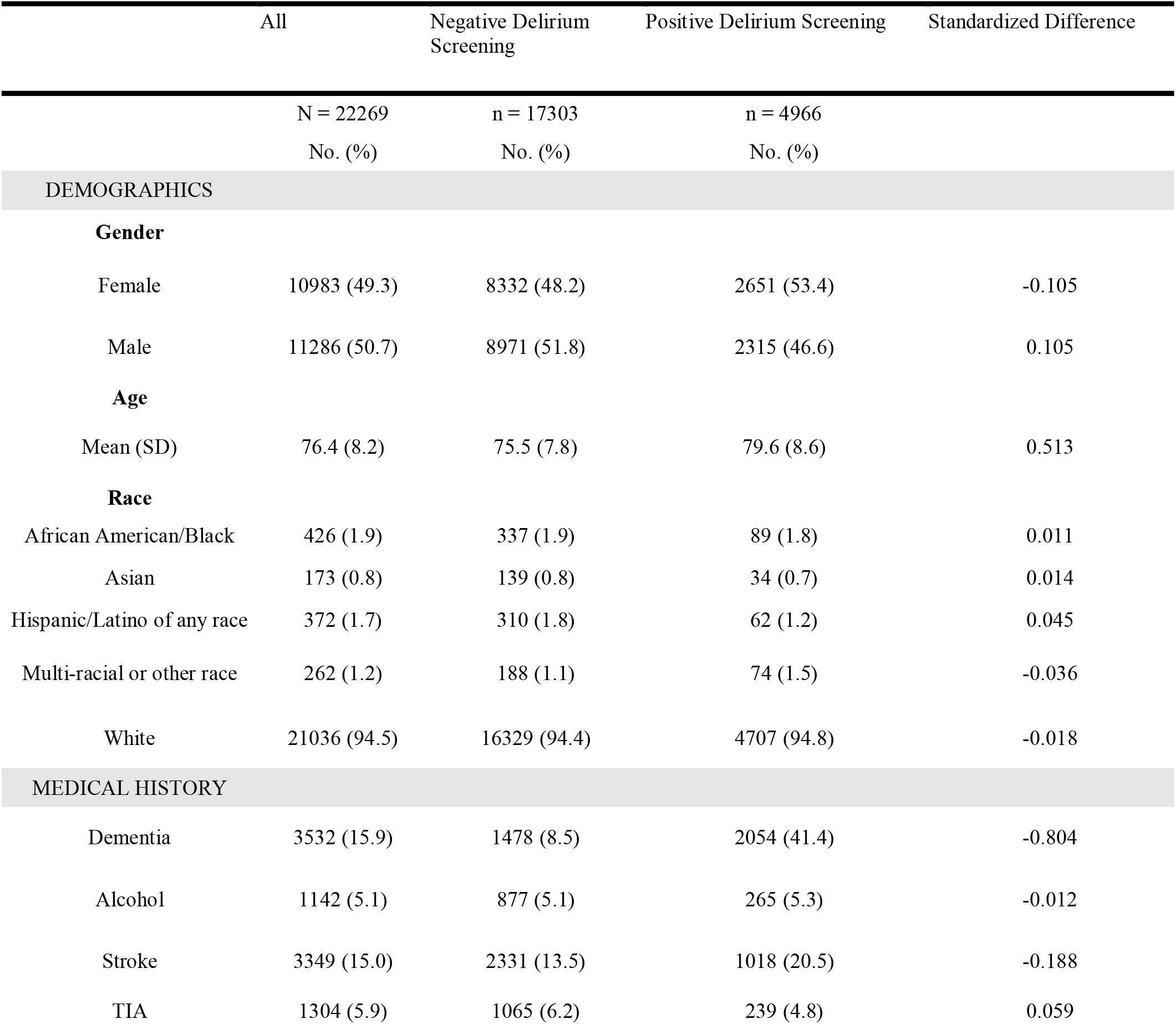
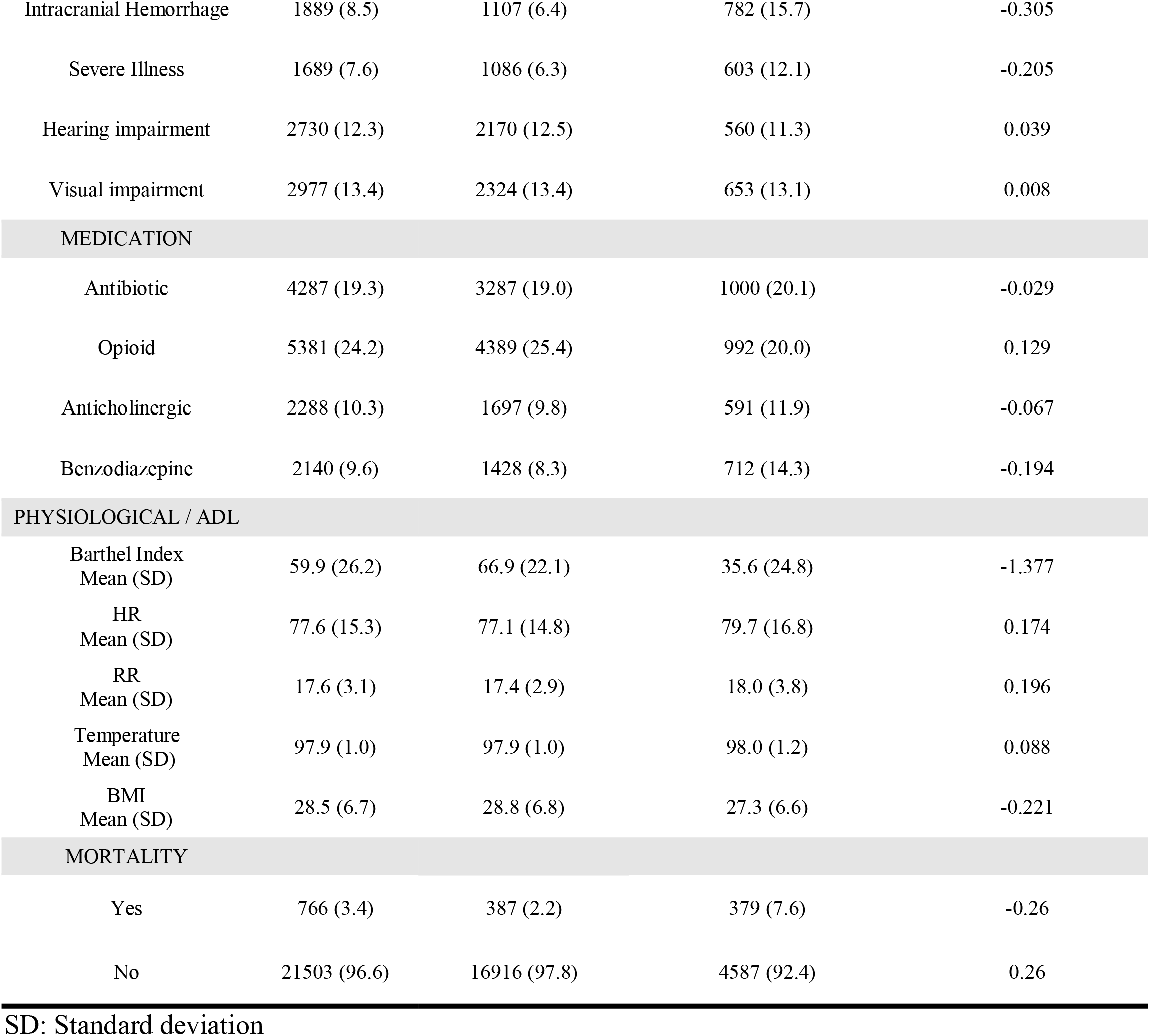
Patient Characteristics.

|  | All | Negative Delirium Screening | Positive Delirium Screening | Standardized Difference |
| --- | --- | --- | --- | --- |
|  | N = 22269 | n = 17303 | n = 4966 |  |
|  | No. (%) | No. (%) | No. (%) |  |
| **DEMOGRAPHICS** |  |  |  |  |
| **Gender** |  |  |  |  |
| Female | 10983 (49.3) | 8332 (48.2) | 2651 (53.4) | -0.105 |
| Male | 11286 (50.7) | 8971 (51.8) | 2315 (46.6) | 0.105 |
| **Age** |  |  |  |  |
| Mean (SD) | 76.4 (8.2) | 75.5 (7.8) | 79.6 (8.6) | 0.513 |
| **Race** |  |  |  |  |
| African American/Black | 426 (1.9) | 337 (1.9) | 89 (1.8) | 0.011 |
| Asian | 173 (0.8) | 139 (0.8) | 34 (0.7) | 0.014 |
| Hispanic/Latino of any race | 372 (1.7) | 310 (1.8) | 62 (1.2) | 0.045 |
| Multi-racial or other race | 262 (1.2) | 188 (1.1) | 74 (1.5) | -0.036 |
| White | 21036 (94.5) | 16329 (94.4) | 4707 (94.8) | -0.018 |
| **MEDICAL HISTORY** |  |  |  |  |
| Dementia | 3532 (15.9) | 1478 (8.5) | 2054 (41.4) | -0.804 |
| Alcohol | 1142 (5.1) | 877 (5.1) | 265 (5.3) | -0.012 |
| Stroke | 3349 (15.0) | 2331 (13.5) | 1018 (20.5) | -0.188 |
| TIA | 1304 (5.9) | 1065 (6.2) | 239 (4.8) | 0.059 |
| Intracranial Hemorrhage | 1889 (8.5) | 1107 (6.4) | 782 (15.7) | -0.305 |
| Severe Illness | 1689 (7.6) | 1086 (6.3) | 603 (12.1) | -0.205 |
| Hearing impairment | 2730 (12.3) | 2170 (12.5) | 560 (11.3) | 0.039 |
| Visual impairment | 2977 (13.4) | 2324 (13.4) | 653 (13.1) | 0.008 |
| MEDICATION |  |  |  |  |
| Antibiotic | 4287 (19.3) | 3287 (19.0) | 1000 (20.1) | -0.029 |
| Opioid | 5381 (24.2) | 4389 (25.4) | 992 (20.0) | 0.129 |
| Anticholinergic | 2288 (10.3) | 1697 (9.8) | 591 (11.9) | -0.067 |
| Benzodiazepine | 2140 (9.6) | 1428 (8.3) | 712 (14.3) | -0.194 |
| PHYSIOLOGICAL / ADL |  |  |  |  |
| Barthel Index<br>Mean (SD) | 59.9 (26.2) | 66.9 (22.1) | 35.6 (24.8) | -1.377 |
| HR<br>Mean (SD) | 77.6 (15.3) | 77.1 (14.8) | 79.7 (16.8) | 0.174 |
| RR<br>Mean (SD) | 17.6 (3.1) | 17.4 (2.9) | 18.0 (3.8) | 0.196 |
| Temperature<br>Mean (SD) | 97.9 (1.0) | 97.9 (1.0) | 98.0 (1.2) | 0.088 |
| BMI<br>Mean (SD) | 28.5 (6.7) | 28.8 (6.8) | 27.3 (6.6) | -0.221 |
| MORTALITY |  |  |  |  |
| Yes | 766 (3.4) | 387 (2.2) | 379 (7.6) | -0.26 |
| No | 21503 (96.6) | 16916 (97.8) | 4587 (92.4) | 0.26 |

SD: Standard deviation

**Figure 1.**
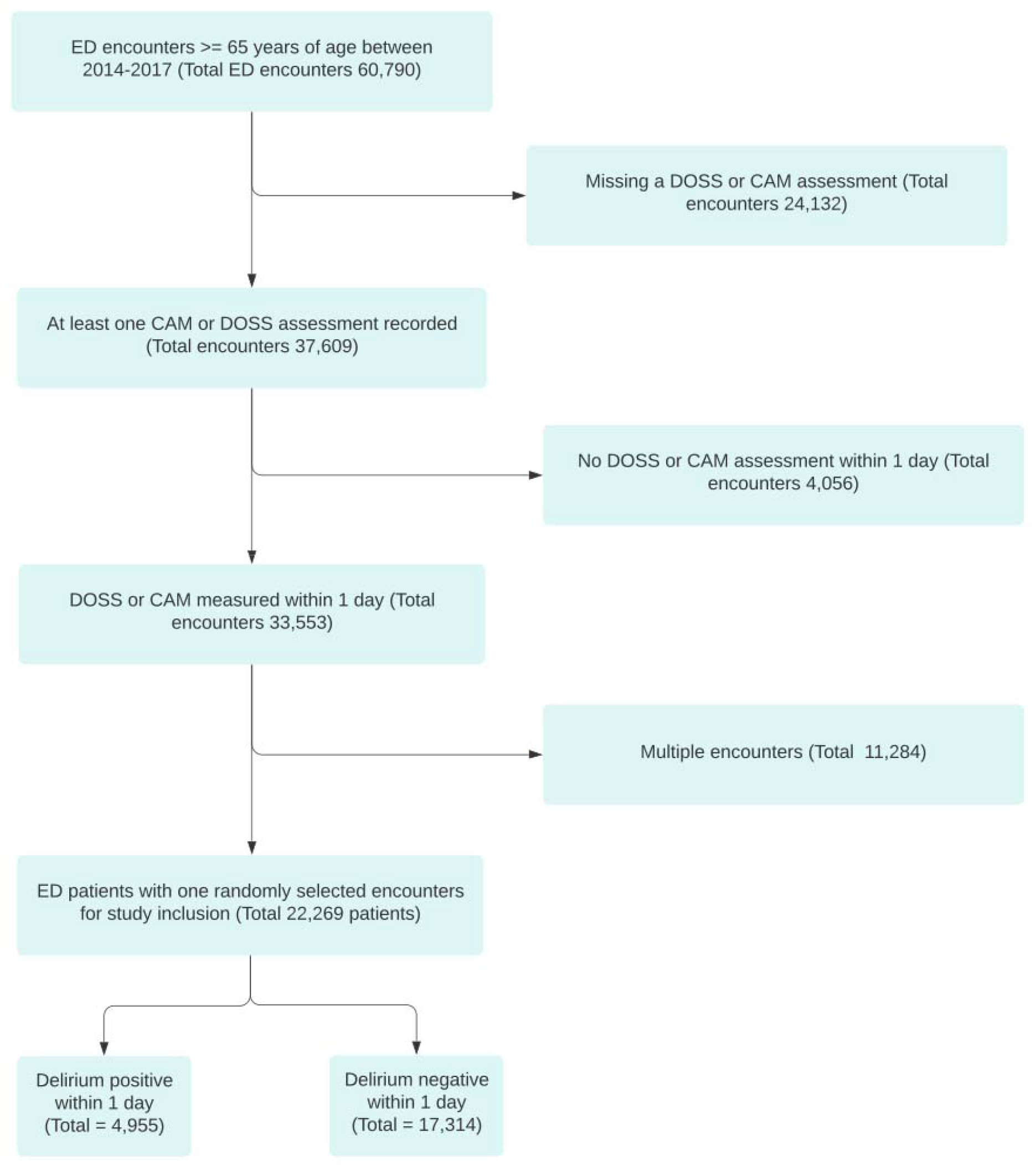
Flow chart of patient selection.

The mean age was 76.4 (SD 8.2), and 50.7% were male. A total of 21,036 (94.5%) were white. The mean BMI was 28.5 (SD 6.7). Overall, 3,532 (15.9%) of them had a past history of dementia, 3,349 (15.0%) had a past history of stroke, and 1,889 (8.5%) had a diagnosis of intracranial hemorrhage in the ED. The rest of the variables, vital signs, and medications that were given in the ED are shown in Table 1. Predictors with missing values were respiratory rate (0.3%), heart rate (0.3%), BMI (2.4%), and the Barthel index (2.7%). Lastly, a total of 766 (3.4%) died during hospitalization.

### Model Specification

We reported the top 3 of the full prediction models to allow predictions for individuals. Tables 2, 3, and 4 have a list of variables used in the logistic regression, random forest, and gradient boosting machine models. These tables list coefficients for logistic regression and variable importance for the two other models (Tables 2, 3, and 4).

**Table 2.** Logistic Regression.

| Feature | Coefficient | Coefficient 95% CI | Odds Ratio | Odds Ratio 95% CI |
| --- | --- | --- | --- | --- |
| Dementia | 1.88 | (1.80, 1.96) | 6.53 | (6.02, 7.09) |
| Intracranial Hemorrhage | 1.01 | (0.90, 1.12) | 2.75 | (2.47, 3.07) |
| Benzodiazepine | 0.69 | (0.58, 0.80) | 1.99 | (1.78, 2.22) |
| Stroke | 0.50 | (0.41, 0.59) | 1.65 | (1.50, 1.80) |
| Severe Illness | 0.48 | (0.35, 0.62) | 1.62 | (1.42, 1.85) |
| Age | 0.33 | (0.30, 0.37) | 1.39 | (1.34, 1.45) |
| Alcohol | 0.21 | (0.06, 0.37) | 1.24 | (1.06, 1.45) |
| Antibiotic | 0.16 | (0.07, 0.25) | 1.17 | (1.07, 1.28) |
| Anticholinergic | 0.14 | (0.02, 0.25) | 1.15 | (1.02, 1.29) |
| HR | 0.12 | (0.08, 0.16) | 1.13 | (1.09, 1.17) |
| RR | 0.11 | (0.08, 0.15) | 1.12 | (1.08, 1.16) |
| BMI | -0.08 | (-0.11, -0.04) | 0.93 | (0.89, 0.96) |
| Intercept | -2.08 | (-2.13, -2.02) | 0.12 | (0.12, 0.13) |

**Table 3.** Random Forest.

| Feature | Variable Importance |
| --- | --- |
| Dementia | 100.0 |
| Age | 42.7 |
| BMI | 37.7 |
| HR | 30.3 |
| RR | 26.0 |
| Intracranial Hemorrhage | 15.2 |
| Stroke | 8.8 |
| Benzodiazepine | 8.5 |
| Severe Illness | 6.2 |
| Antibiotic | 4.0 |
| Anticholinergic | 3.5 |
| Alcohol | 2.8 |

Variable importance relative to the feature with the highest importance on a 0-100 scale.

**Table 4.** GBM.

| Feature | Variable Importance |
| --- | --- |
| Dementia | 100.0 |
| Age | 51.0 |
| BMI | 51.0 |
| HR | 40.5 |
| RR | 35.6 |
| Intracranial Hemorrhage | 17.1 |
| Benzodiazepine | 10.7 |
| Stroke | 10.3 |
| Severe Illness | 6.7 |
| Antibiotic | 5.1 |
| Anticholinergic | 5.1 |
| Alcohol | 4.6 |

Variable importance relative to the feature with the highest importance on a 0-100 scale.

Table 2 outlines the coefficients for the logistic regression model with an inverse regularization parameter of .5. Relative feature importance scores were similar for the gradient-boosted machine and random forest models (Tables 2, 3, and 4). Highly ranked variables included the history of dementia, age, and BMI. The Barthel Index score also highly ranked when it was included. (Supplemental table 2, 3, and 4)

These prediction models were developed to put variables into a model-based probability calculator. This practice can be implemented in any electronic medical record using an equation or an online calculator. For example, we estimated the probability of a positive delirium screening based on the combination of demographic and clinical data (Table 5).

**Table 5.** The probabilities of positive delirium screen for selected case examples.

| Cases | Logistic regression | Random forest | Gradient boost model |
| --- | --- | --- | --- |
| Case 1. Age 70, history of dementia, intracranial hemorrhage (ICH), stroke, severe illness, alcohol, use of benzodiazepines, anticholinergics, antibiotics, and respiratory rate (RR) 24, heart rate (HR) 110, body mass index (BMI) 30. | 96% | 65% | 74% |
| Case 2. Age 65, No dementia, no ICH, no stroke, no severe illness, no alcohol, and no use of benzodiazepines, anticholinergics, or antibiotics, and RR 20, HR 60, and BMI 25 | 7% | 10% | 9% |
| Case 3. Age 70, No dementia, no ICH, with history of stroke, and severe illness, without benzodiazepines, anticholinergics, and used antibiotics, with RR 24, HR 100, BMI 28. | 31% | 28% | 29% |

### Model Performance

Figures 2 and 3 illustrate the predictive performance of each model with receiver operating characteristic (ROC) curves (Figure 2, 3). The area under the ROC curves ranged from .69 to .77. In this analysis, RF and GBM followed by LR demonstrated the best predictive ability with respective AUCs of .772 (95% CI .77, .774), .77 (95% CI .768, .772), and .767 (95%CI .764, .769). At the 90% sensitivity threshold, RF, GBM and LR models achieved a specificity of 33-35%. When the Barthel index was added to the model, we noticed improved accuracy and AUC (Table 6).

**Table 6.**
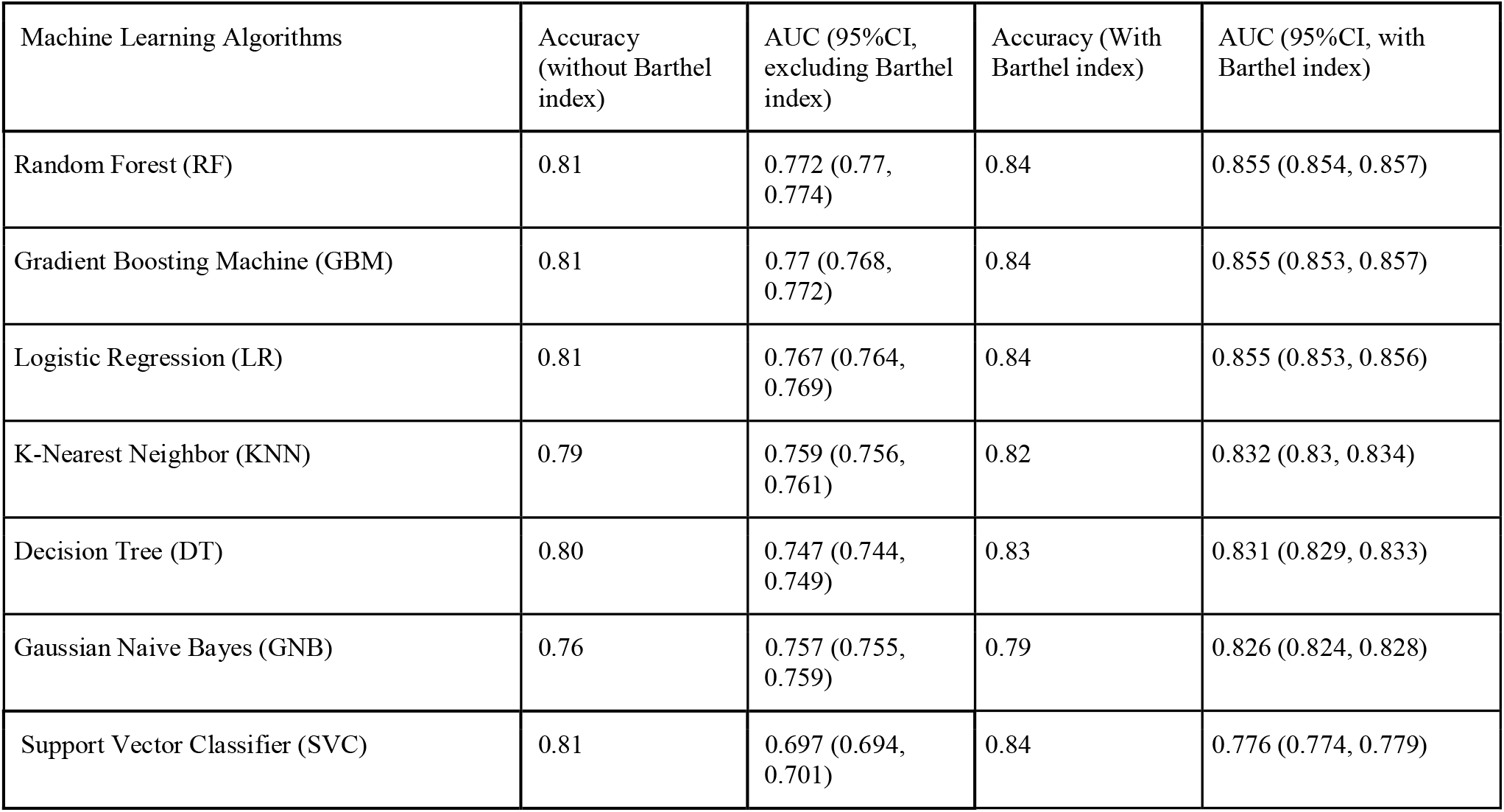
Diagnostic characteristics for machine learning models.

| Machine Learning Algorithms | Accuracy<br>(without Barthel<br>index) | AUC (95%CI,<br>excluding Barthel<br>index) | Accuracy (With<br>Barthel index) | AUC (95%CI, with<br>Barthel index) |
| --- | --- | --- | --- | --- |
| Random Forest (RF) | 0.81 | 0.772 (0.77,<br>0.774) | 0.84 | 0.855 (0.854, 0.857) |
| Gradient Boosting Machine (GBM) | 0.81 | 0.77 (0.768,<br>0.772) | 0.84 | 0.855 (0.853, 0.857) |
| Logistic Regression (LR) | 0.81 | 0.767 (0.764,<br>0.769) | 0.84 | 0.855 (0.853, 0.856) |
| K-Nearest Neighbor (KNN) | 0.79 | 0.759 (0.756,<br>0.761) | 0.82 | 0.832 (0.83, 0.834) |
| Decision Tree (DT) | 0.80 | 0.747 (0.744,<br>0.749) | 0.83 | 0.831 (0.829, 0.833) |
| Gaussian Naive Bayes (GNB) | 0.76 | 0.757 (0.755,<br>0.759) | 0.79 | 0.826 (0.824, 0.828) |
| Support Vector Classifier (SVC) | 0.81 | 0.697 (0.694,<br>0.701) | 0.84 | 0.776 (0.774, 0.779) |

**Figure 2.**
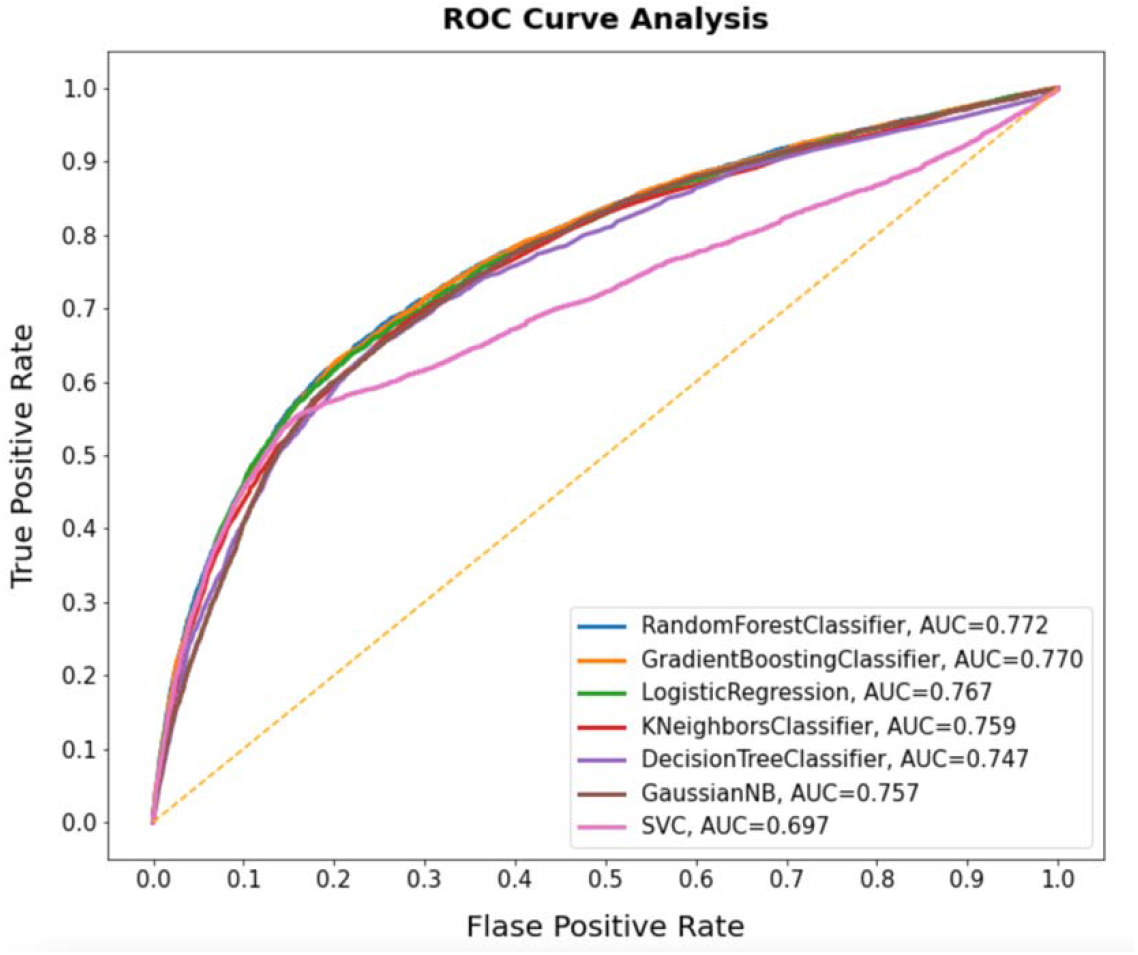
AUC curves for the machine learning algorithms (without Barthel index)

**Figure 3.**
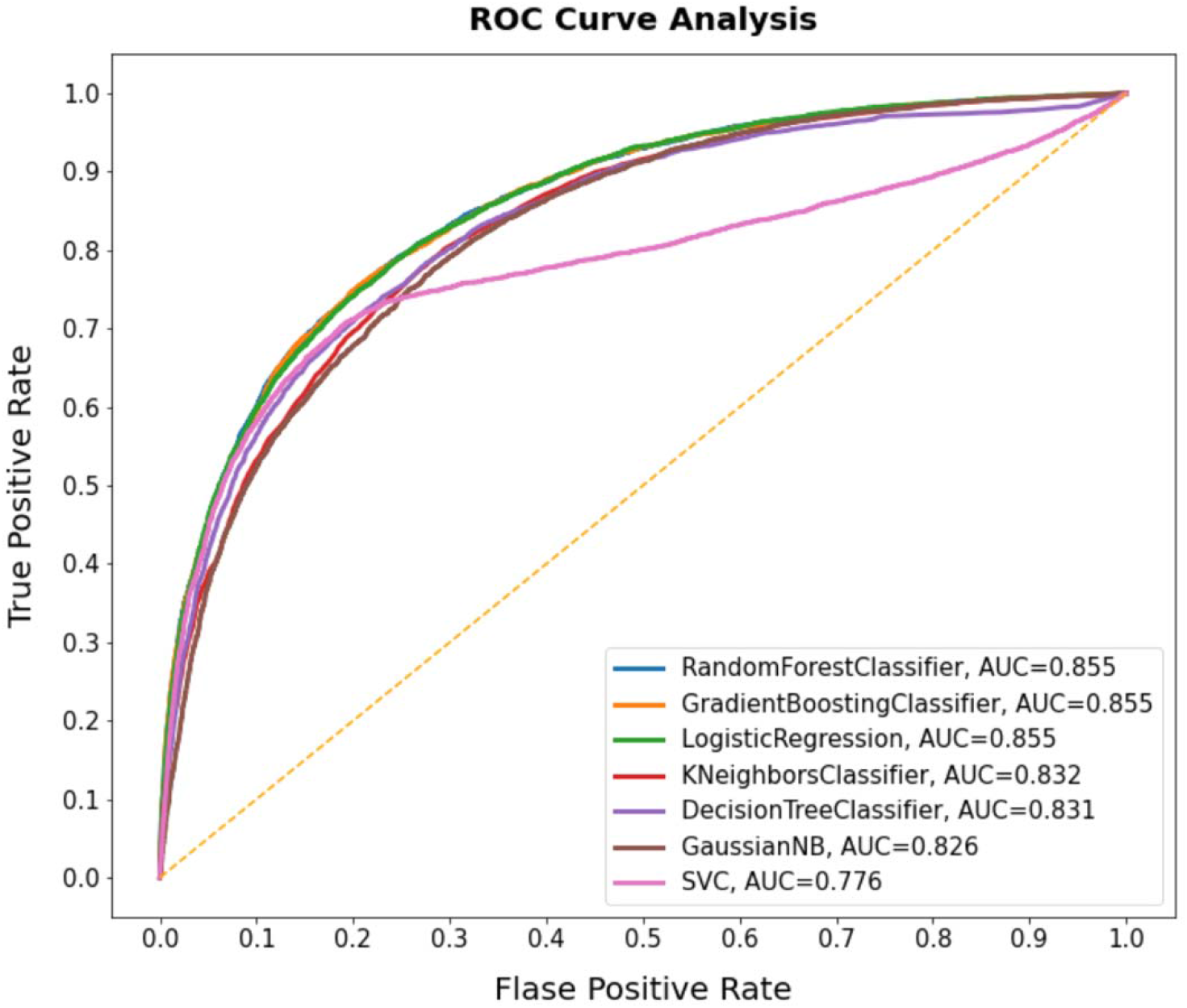
AUC curves for the machine learning algorithms (with Barthel index)

## DISCUSSION

This study used several variables from the ED to develop and validate the delirium prediction model within the first 24 hours of hospitalization. The rate of delirium based on the screening was up to 21% within the first 24 hours, which underscores the need to screen for delirium early in the hospital stay, preferably in the ED, and prevent or treat delirium as early as possible. Our study identified a combination of variables that included demographic information, medical history, and medications by using a machine-learning approach to predict delirium. We also explored use of the Barthel index, a measure of functional status, which was measured after hospital admission, since that could inform delirium risk after hospitalization.

We identified that the rate of delirium based on the screening was up to 21% within the first 24 hours. The prevalence of delirium in the emergency department (ED) is estimated to be 8%–17%, but it is difficult to identify without a screening process.^1-3^ The lack of an effective screening process leads to underdiagnosed and undertreated delirium.^5^ The prevalence of delirium in the inpatient unit, including intensive care units, is 18%–64% in the literature, which underscores the need for prevention and treatment strategy prior to hospitalization.^19^ Although several cognitive assessment tools exist, they require training and dissemination, and compliance can be limited without strong merit and support from leadership and stakeholders.^6-8^ Early screening and interventional options are emerging and seem promising, as reported by several recent studies, but these programs depend on effective screening.^9-11^ Our study may help to narrow down the high-risk patient population who need further delirium screening.

Our study identified delirium using DOSS and CAM-ICU as the reference standards in the clinical setting. The fluctuating nature of delirium poses a challenge to clinicians, as evidenced by Lewis et al., who reported that the estimated misdiagnosis rate of delirium in the ED was up to 80% in the 1990s, and the rate of misdiagnosis has remained high.^5,16^ The study’s findings highlight the limitation of clinical gestalt without any additional diagnostic modality or assessment tools. Our inpatient unit routinely uses the DOSS and CAM-ICU for ventilated patients, and validation studies showed that sensitivity and specificity were above 90%.^20,21^ Because of their superior accuracy over clinical gestalt in the ED, we used DOSS and CAM-ICU as an approximation of the delirium outcome in this study. This approach likely included both ones who presented with delirium in the ED and ones who developed delirium after ED visit. A future study should investigate both the prevalence and the incidence of delirium by screening for delirium in the ED and inpatient unit to explore the distinctive features of those who come to the ED with delirium and those who develop delirium during a hospital stay.

The Area Under Curve (AUC), sensitivity, specificity for the RF, GBM, and LR were 0.76-0.77, 0.90, 0.33-0.35, respectively. These improved significantly after the Barthel index was included (Table 6). The probability of developing delirium based on the three scenarios ranged from 7% to 96% (Table 5). We previously analyzed the diagnostic characteristics of three delirium prediction models that could be applied in the past: delirium risk score, risk prediction model, and susceptibility score.^3,16,22^ These models were examined in our retrospective hospital-wide data, and AUC ranged from 0.71 to 0.8.^23^ The use of two delirium screenings, the combination of variables, and the machine learning approach improved our model’s predictive ability only after the Barthel index was included. Our model prioritized higher sensitivity as it can be used as a screening tool, possibly triggered by electronic health records. It requires confirmatory tests for delirium.

Our study reported the importance of variables, such as age and dementia which cannot be modified, heart rate, respiratory rate, and severe illness, which can be modified but may reflect underlying illness which may or may not be modifiable. The Barthel index is an important variable that is potentially modifiable depending on the cause of functional deficits, for example with feeding assistance and early mobilization, but may also reflect the effects of delirium. The importance of benzodiazepines and anticholinergics was not high, but these drugs can be decreased with a deprescribing program.^24,25^ These findings highlighted several modifiable variables that we can approach in the ED and hospital setting.

Our prediction models were developed by machine learning algorithms. We selected random forest, gradient boosting machine, logistic regression, KNN, decision tree, Gaussian Naïve Bayes, and SVC. The use of GBM, RF demonstrated the highest AUC, and this was similar to the logistic regression-based model. The recent study of the prediction of postoperative delirium by Wong et al. used penalized logistic regression, GBM, artificial neural network with a single hidden layer, and linear support vector machine, and the random forest and the GBM model reported an AUC of 0.855.^26^ The comparison of the machine learning-driven models added a significant impact on how to identify patients who have a high risk of developing delirium while minimizing bias.

### Strengths and Limitations

The strength of our study was that we used either DOSS or CAM-ICU on all hospitalized older adults, so the study was able to provide a large dataset to derive and validate the delirium prediction model with minimal variance. We used the outcome of positive delirium screening in the first 24 hours as the best available proxy for delirium in the ED. Since delirium is missed in the ED and our prediction model will help to identify a high-risk group, which fits with using the model to identify prevalent and impending delirium. Another strength is that the use of interpretable machine learning algorithms with cross-validation enabled us to identify the best model with minimal bias and avoid overfitting. Lastly, the list of probability with the presence or absence of predictors will be informative to clinicians utilizing the tool in the clinical setting.

There are several limitations that are important to recognize. First, our institution is unique in that routine delirium screening is available, but almost half of the admitted older adults did not have delirium screening within the first 24 hours of hospitalization, so it is prone to measurement bias. Second, we took the first Barthel index in the inpatient unit, so it may have measured the functional status after they developed delirium. We thought that Barthel index was unlikely to change within the 24 hours of hospitalization following ED arrival, but reverse causality from the effects of delirium on functional status is possible. The model without the Barthel index still demonstrated accuracy for delirium, but we explored use of the Barthel index in the model since the loss of activity of daily living (ADL) is an important predictor for delirium, and ADL can be estimated in the ED.^16^ Third, a list of anticholinergics is vast, and the drug flag may not capture all anticholinergics. We reviewed drug names under anticholinergics several times and updated data to overcome this challenge. Fourth, this study was conducted in a primarily white population and would benefit from further validation in a more diverse population.

## CONCLUSION

This study demonstrated the use of machine learning algorithms to identify the combination of variables that are predictive of delirium within the 24 hours of hospitalization from the ED. The discovery of a predictive model that clinicians can use as a clinical decision aid could lead to improved detection of delirium and identification of a high-risk group. This contribution is significant because the findings will introduce a clinical decision aid that either clinicians use actively or receive passively from machine learning algorithms, overcoming the limitation of misdiagnosis or under diagnosis by clinical gestalt alone to detect delirium. Our future objective will be to develop a clinical decision aid integrated into electronic medical record to predict delirium in real-time, so ED providers and the inpatient team can focus on delirium screening for high-risk individuals and implement a delirium prevention program.

## Data Availability

Data files are available in GitHub as listed below.

https://github.com/briannamueller/Delirium-Analyisis

## Acknowledgement

This study was supported by an award to Sangil Lee by the NIDUS delirium network (No. NIA R24AG054259). Research reported in this publication was supported by the National Center For Advancing Translational Sciences of the National Institutes of Health under Award Number UL1TR002537. The content is solely the responsibility of the authors and does not necessarily represent the official views of the National Institutes of Health

---

**Supplemental table 1.** Anticholinergics Received by the Sample (generic name, frequency and percentage)

| Generic Name | Frequency | Percent |
| --- | --- | --- |
| Amitriptyline 10 mg Tablet | 61 | 0.77 |
| Amitriptyline 100 mg Tablet | 7 | 0.09 |
| Amitriptyline 150 mg Tablet | 3 | 0.04 |
| Amitriptyline 25 mg Tablet | 75 | 0.94 |
| Amitriptyline 50 mg Tablet | 53 | 0.67 |
| Amitriptyline 75 mg Tablet | 11 | 0.14 |
| Amitriptyline Oral | 5 | 0.06 |
| Atropine 0.1 mg/ml Injection Syringe | 86 | 1.08 |
| Atropine 0.4 mg/ml Injection Solution | 65 | 0.82 |
| Atropine Oral | 1 | 0.01 |
| Belladonna Alkaloids-Opium 16.2 mg-30 mg Rectal Suppository | 3 | 0.04 |
| Belladonna Alkaloids-Opium 16.2 mg-60 mg Rectal Suppository | 33 | 0.41 |
| Benztropine 0.5 mg Tablet | 25 | 0.31 |
| Benztropine 1 mg Tablet | 21 | 0.26 |
| Benztropine 1 mg/ml Injection Solution | 23 | 0.29 |
| Benztropine 2 mg Tablet | 8 | 0.1 |
| Carbinoxamine-Pseudoephedrine Oral | 1 | 0.01 |
| Chlordiazepoxide-Clidinium 5 mg-2.5 mg Capsule | 5 | 0.06 |
| Chlorpheniramine 2 mg-Phenylephrine 5 mg-Acetaminophen 325 mg Tablet | 3 | 0.04 |
| Chlorpheniramine 4 mg Tablet | 6 | 0.08 |
| Chlorpheniramine Oral | 1 | 0.01 |
| Chlorpheniramine-Phenylephrine-Acetaminophen Oral | 1 | 0.01 |
| Chlorpheniramine-Pseudoephedrine Oral | 1 | 0.01 |
| Chlorpheniramine-Pseudoephedrine-Acetaminophen Oral | 1 | 0.01 |
| Chlorpheniramine-Pseudoephedrine-Ibuprofen Oral | 2 | 0.03 |
| Chlorpromazine 10 mg Tablet | 2 | 0.03 |
| Chlorpromazine 100 mg Tablet | 5 | 0.06 |
| Chlorpromazine 25 mg Tablet | 5 | 0.06 |
| Chlorpromazine 25 mg/ml Injection Solution | 4 | 0.05 |
| Chlorpromazine 50 mg Tablet | 4 | 0.05 |
| Cimetidine 200 mg Tablet | 3 | 0.04 |
| Clemastine 1.34 mg Tablet | 1 | 0.01 |
| Clomipramine 50 mg Capsule | 2 | 0.03 |
| Clozapine 100 mg Tablet | 34 | 0.43 |
| Clozapine 200 mg Tablet | 1 | 0.01 |
| Clozapine 25 mg Tablet | 1 | 0.01 |
| Clozapine 50 mg Tablet | 1 | 0.01 |
| Cyclobenzaprine 10 mg Tablet | 550 | 6.91 |
| Cyclobenzaprine 5 mg Tablet | 100 | 1.26 |
| Cyclobenzaprine Oral | 3 | 0.04 |
| Cyproheptadine 4 mg Tablet | 4 | 0.05 |
| Darifenacin ER 15 mg Tablet, Extended Release 24 Hr | 2 | 0.03 |
| Desipramine 10 mg Tablet | 1 | 0.01 |
| Desipramine 100 mg Tablet | 4 | 0.05 |
| Desipramine 25 mg Tablet | 2 | 0.03 |
| Desipramine 50 mg Tablet | 1 | 0.01 |
| Dicyclomine 10 mg Capsule | 47 | 0.59 |
| Dicyclomine 10 mg/5 ml Oral Solution | 1 | 0.01 |
| Dicyclomine 10 mg/ml Intramuscular Solution | 53 | 0.67 |
| Dicyclomine 20 mg Tablet | 79 | 0.99 |
| Dicyclomine Oral | 2 | 0.03 |
| Dimenhydrinate 50 mg Tablet | 3 | 0.04 |
| Diphenhydramine 12.5 mg Chewable Tablet | 1 | 0.01 |
| Diphenhydramine 12.5 mg/5 ml Oral Elixir | 24 | 0.3 |
| Diphenhydramine 12.5 mg/5 ml Oral Liquid | 9 | 0.11 |
| Diphenhydramine 25 mg Capsule | 707 | 8.88 |
| Diphenhydramine 25 mg Tablet | 11 | 0.14 |
| Diphenhydramine 25 mg-Acetaminophen 500 mg Tablet | 52 | 0.65 |
| Diphenhydramine 25 mg-Acetaminophen 500 mg/15 ml Oral Solution | 1 | 0.01 |
| Diphenhydramine 50 mg Capsule | 30 | 0.38 |
| Diphenhydramine 50 mg/ml Injection Solution | 1170 | 14.69 |
| Diphenhydramine Oral | 7 | 0.09 |
| Diphenhydramine-Acetaminophen Oral | 12 | 0.15 |
| Diphenhydramine-Phenylephrine-Acetaminophen-Guaifenesin Oral | 1 | 0.01 |
| Diphenoxylate-Atropine 2.5 mg-0.025 mg Tablet | 59 | 0.74 |
| Diphenoxylate-Atropine 2.5 mg-0.025 mg/5 ml Oral Liquid | 1 | 0.01 |
| Doxepin 10 mg Capsule | 11 | 0.14 |
| Doxepin 10 mg/ml Oral Concentrate | 1 | 0.01 |
| Doxepin 100 mg Capsule | 7 | 0.09 |
| Doxepin 150 mg Capsule | 1 | 0.01 |
| Doxepin 25 mg Capsule | 8 | 0.1 |
| Doxepin 50 mg Capsule | 5 | 0.06 |
| Doxepin Oral | 1 | 0.01 |
| Doxylamine Succinate 25 mg Tablet | 5 | 0.06 |
| Doxylamine Succinate Oral | 1 | 0.01 |
| Doxylamine-Pseudoephedrine-Dextromethorphan-Acetaminophen Oral | 2 | 0.03 |
| Fesoterodine ER 4 mg Tablet, Extended Release 24 Hr | 9 | 0.11 |
| Fesoterodine ER 8 mg Tablet, Extended Release 24 Hr | 2 | 0.03 |
| Flavoxate 100 mg Tablet | 1 | 0.01 |
| Glycopyrrolate 0.2 mg/ml Injection Solution | 211 | 2.65 |
| Glycopyrrolate 1 mg Tablet | 8 | 0.1 |
| Glycopyrrolate 1 mg/5 ml (0.2 mg/ml) Oral Solution | 9 | 0.11 |
| Glycopyrrolate 2 mg Tablet | 2 | 0.03 |
| Hydrocodone 10 mg-Chlorpheniramine 8 mg/5 ml Oral Susp Extend. Rel 12Hr | 2 | 0.03 |
| Hydrocodone-Homatropine 5 mg-1.5 mg Tablet | 1 | 0.01 |
| Hydrocodone-Homatropine 5 mg-1.5 mg/5 ml Oral Syrup | 1 | 0.01 |
| Hydroxyzine HCl 10 mg Tablet | 31 | 0.39 |
| Hydroxyzine HCl 10 mg/5 ml Oral Solution | 3 | 0.04 |
| Hydroxyzine HCl 25 mg Tablet | 334 | 4.19 |
| Hydroxyzine HCl 50 mg Tablet | 31 | 0.39 |
| Hydroxyzine HCl 50 mg/ml Intramuscular Solution | 9 | 0.11 |
| Hydroxyzine HCl Oral | 2 | 0.03 |
| Hydroxyzine Pamoate 100 mg Capsule | 1 | 0.01 |
| Hydroxyzine Pamoate 25 mg Capsule | 102 | 1.28 |
| Hydroxyzine Pamoate 50 mg Capsule | 8 | 0.1 |
| Hyoscyamine 0.125 mg Sublingual Tablet | 1 | 0.01 |
| Hyoscyamine ER 0.375 mg Tablet, Extended Release,12 Hr | 1 | 0.01 |
| Hyoscyamine Sulfate 0.125 mg Tablet | 7 | 0.09 |
| Hyoscyamine Sulfate Oral | 2 | 0.03 |
| Ibuprofen 200 mg-Diphenhydramine HCl 25 mg Capsule | 3 | 0.04 |
| Ibuprofen-Diphenhydramine Citrate 200 mg-38 mg Tablet | 2 | 0.03 |
| Ibuprofen-Diphenhydramine HCl Oral | 3 | 0.04 |
| Imipramine 10 mg Tablet | 1 | 0.01 |
| Imipramine 25 mg Tablet | 7 | 0.09 |
| Imipramine 50 mg Tablet | 9 | 0.11 |
| Imipramine Oral | 1 | 0.01 |
| Ketamine 10 mg/ml Injection Solution | 345 | 4.33 |
| Ketamine 100 mg/ml Injection Solution | 2 | 0.03 |
| Ketamine 50 mg/ml Injection Solution | 27 | 0.34 |
| Loxapine Succinate 10 mg Capsule | 1 | 0.01 |
| Meclizine 12.5 mg Tablet | 82 | 1.03 |
| Meclizine 25 mg Chewable Tablet | 1 | 0.01 |
| Meclizine 25 mg Tablet | 403 | 5.06 |
| Meclizine Oral | 3 | 0.04 |
| Naproxen 220 mg-Diphenhydramine 25 mg Tablet | 1 | 0.01 |
| Nortriptyline 10 mg Capsule | 43 | 0.54 |
| Nortriptyline 25 mg Capsule | 33 | 0.41 |
| Nortriptyline 50 mg Capsule | 38 | 0.48 |
| Nortriptyline 75 mg Capsule | 4 | 0.05 |
| Nortriptyline Oral | 1 | 0.01 |
| Olanzapine 10 mg Disintegrating Tablet | 15 | 0.19 |
| Olanzapine 10 mg Intramuscular Solution | 125 | 1.57 |
| Olanzapine 10 mg Tablet | 21 | 0.26 |
| Olanzapine 15 mg Tablet | 7 | 0.09 |
| Olanzapine 2.5 mg Tablet | 32 | 0.4 |
| Olanzapine 20 mg Tablet | 9 | 0.11 |
| Olanzapine 5 mg Disintegrating Tablet | 42 | 0.53 |
| Olanzapine 5 mg Tablet | 52 | 0.65 |
| Olanzapine 7.5 mg Tablet | 5 | 0.06 |
| Orphenadrine Citrate ER 100 mg Tablet, Extended Release | 2 | 0.03 |
| Orphenadrine Citrate Oral | 2 | 0.03 |
| Oxybutynin 3.9 mg/24 Hr Semiweekly Transdermal Patch | 2 | 0.03 |
| Oxybutynin Chloride 5 mg Tab | 2 | 0.03 |
| Oxybutynin Chloride 5 mg Tablet | 301 | 3.78 |
| Oxybutynin Chloride 5 mg/5 ml Oral Syrup | 2 | 0.03 |
| Oxybutynin Chloride ER 10 mg Tablet, Extended Release 24 Hr | 102 | 1.28 |
| Oxybutynin Chloride ER 15 mg Tablet, Extended Release 24 Hr | 17 | 0.21 |
| Oxybutynin Chloride ER 5 mg Tablet, Extended Release 24 Hr | 56 | 0.7 |
| Oxybutynin Chloride Oral | 7 | 0.09 |
| Paroxetine 10 mg Tablet | 69 | 0.87 |
| Paroxetine 20 mg Tablet | 105 | 1.32 |
| Paroxetine 30 mg Tablet | 39 | 0.49 |
| Paroxetine 40 mg Tablet | 50 | 0.63 |
| Paroxetine ER 12.5 mg Tablet, Extended Release 24 Hr | 1 | 0.01 |
| Paroxetine ER 25 mg Tablet, Extended Release 24 Hr | 1 | 0.01 |
| Paroxetine Oral | 3 | 0.04 |
| Phenobarbital-Hyoscyamine-Atropine-Scopolamine Oral | 1 | 0.01 |
| Procyclidine Oral | 1 | 0.01 |
| Promethazine 12.5 mg Rectal Suppository | 6 | 0.08 |
| Promethazine 12.5 mg Tablet | 30 | 0.38 |
| Promethazine 25 mg Rectal Suppository | 15 | 0.19 |
| Promethazine 25 mg Tablet | 116 | 1.46 |
| Promethazine 25 mg/ml Injection Solution | 83 | 1.04 |
| Promethazine 50 mg Tablet | 1 | 0.01 |
| Promethazine 6.25 mg/5 ml Oral Syrup | 4 | 0.05 |
| Promethazine 6.25 mg-Codeine 10 mg/5 ml Syrup | 85 | 1.07 |
| Promethazine Oral | 2 | 0.03 |
| Promethazine-Dm 6.25 mg-15 mg/5 ml Oral Syrup | 1 | 0.01 |
| Propantheline 15 mg Tablet | 1 | 0.01 |
| Quetiapine 100 mg Tablet | 49 | 0.62 |
| Quetiapine 200 mg Tablet | 34 | 0.43 |
| Quetiapine 25 mg Tab | 30 | 0.38 |
| Quetiapine 25 mg Tablet | 173 | 2.17 |
| Quetiapine 300 mg Tablet | 6 | 0.08 |
| Quetiapine 400 mg Tablet | 11 | 0.14 |
| Quetiapine 50 mg Tablet | 109 | 1.37 |
| Quetiapine ER 150 mg Tablet, Extended Release 24 Hr | 1 | 0.01 |
| Quetiapine ER 300 mg Tablet, Extended Release 24 Hr | 3 | 0.04 |
| Quetiapine ER 400 mg Tablet, Extended Release 24 Hr | 2 | 0.03 |
| Quetiapine ER 50 mg Tablet, Extended Release 24 Hr | 1 | 0.01 |
| Quetiapine Oral | 2 | 0.03 |
| Ranitidine 15 mg/ml Oral Syrup | 6 | 0.08 |
| Ranitidine 150 mg Capsule | 4 | 0.05 |
| Ranitidine 150 mg Tablet | 253 | 3.18 |
| Ranitidine 300 mg Capsule | 2 | 0.03 |
| Ranitidine 300 mg Tablet | 49 | 0.62 |
| Ranitidine 75 mg Tablet | 21 | 0.26 |
| Ranitidine Oral | 17 | 0.21 |
| Scopolamine 0.4 mg/ml Injection Solution | 1 | 0.01 |
| Scopolamine 1 mg Over 3 Days Transdermal Patch | 27 | 0.34 |
| Solifenacin 10 mg Tablet | 20 | 0.25 |
| Solifenacin 5 mg Tablet | 40 | 0.5 |
| Thioridazine 25 mg Tablet | 1 | 0.01 |
| Tolterodine 2 mg Tablet | 6 | 0.08 |
| Tolterodine ER 2 mg Capsule, Extended Release 24 Hr | 11 | 0.14 |
| Tolterodine ER 4 mg Capsule, Extended Release 24 Hr | 46 | 0.58 |
| Trifluoperazine 5 mg Tablet | 2 | 0.03 |
| Trihexyphenidyl 2 mg Tablet | 11 | 0.14 |
| Trihexyphenidyl 5 mg Tablet | 1 | 0.01 |
| Tripolidine-Pseudoephedrine 2.5 mg-60 mg Tablet | 1 | 0.01 |
| Trospium 20 mg Tablet | 8 | 0.1 |

**Supplemental figure 1.**
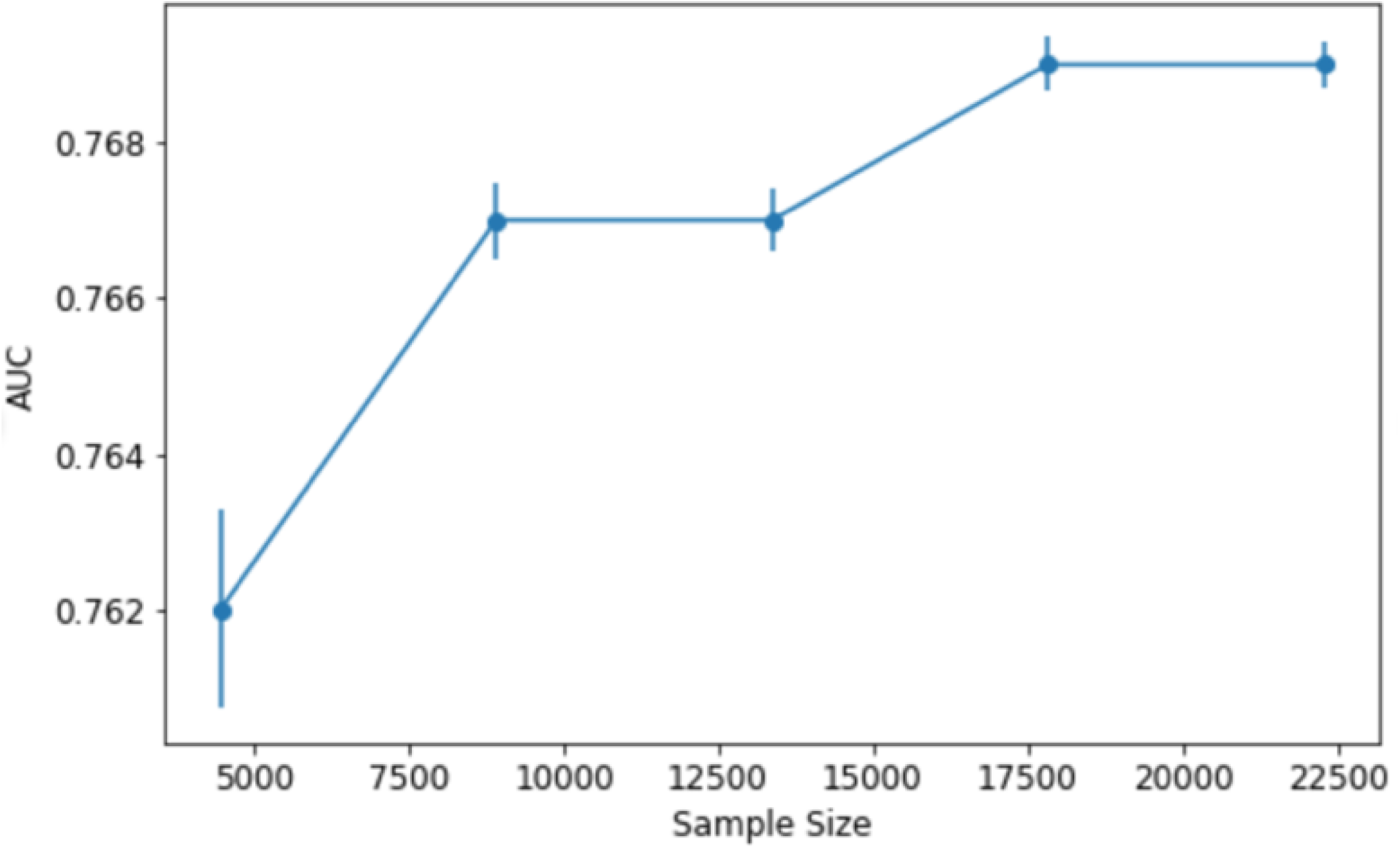
Sample size plot

**Supplemental Table 2.** Logistic regression including Barthel index

| Feature | Coefficient | Coefficient 95% CI | Odds Ratio | Odds Ratio 95% CI |
| --- | --- | --- | --- | --- |
| Dementia | 1.57 | (1.48, 1.66) | 4.82 | (4.40, 5.28) |
| Benzodiazepine | 0.56 | (0.44, 0.68) | 1.75 | (1.55, 1.98) |
| Intracranial Hemorrhage | 0.55 | (0.43, 0.67) | 1.74 | (1.54,1.96) |
| Stroke | 0.26 | (0.16, 0.37) | 1.30 | (1.18, 1.44) |
| Severe Illness | 0.22 | (0.07, 0.37) | 1.25 | (1.08, 1.45) |
| Alcohol | 0.21 | (0.04, 0.38) | 1.24 | (1.04, 1.47) |
| Age | 0.12 | (0.08, 0.16) | 1.13 | (1.09,1.18) |
| RR | 0.08 | (0.05, 0.12) | 1.09 | (1.05, 1.13) |
| Anticholinergic | 0.04 | (-0.09, 0.17) | 1.04 | (0.92, 1.18) |
| HR | 0.03 | (-0.01, 0.08) | 1.03 | (0.99, 1.08) |
| Antibiotic | 0.00 | (-0.10, 0.10) | 1.00 | (0.91, 1.10) |
| BMI | -0.15 | (-0.19, -0.12) | 0.86 | (0.83, 0.90) |
| Barthel | -1.15 | (-1.19, -1.10) | 0.32 | (0.30, 0.33) |
| Intercept | -2.18 | (-2.24, -2.11) | 0.11 | (0.11,0.12) |

**Supplemental Table 3.** Random Forest including Barthel index

| Feature | Variable Importance |
| --- | --- |
| Barthel Index | 100.0 |
| Dementia | 45.6 |
| Age | 19.5 |
| BMI | 15.5 |
| HR | 15.5 |
| RR | 11.6 |
| Intracranial Hemorrhage | 5.6 |
| Stroke | 3.8 |
| Benzodiazepine | 3.7 |
| Severe Illness | 2.6 |
| Antibiotic | 2.0 |
| Anticholinergic | 1.8 |
| Alcohol | 1.6 |

**Supplemental Table 4.** GBM including Barthel index

| Feature | Variable Importance |
| --- | --- |
| Barthel Index | 100.0 |
| Dementia | 45.5 |
| BMI | 24.3 |
| Age | 20.8 |
| HR | 18.5 |
| RR | 16.5 |
| Intracranial Hemorrhage | 6.5 |
| Benzodiazepine | 4.4 |
| Stroke | 4.1 |
| Severe Illness | 2.9 |
| Alcohol | 2.3 |
| Antibiotic | 2.2 |
| Anticholinergic | 2.1 |

## References

1. Francis J, Martin D, Kapoor WN. A prospective study of delirium in hospitalized elderly. JAMA. 1990;263(8):1097–1101.

2. O’Keeffe S, Lavan J. The prognostic significance of delirium in older hospital patients. J Am Geriatr Soc. 1997;45(2):174–178.

3. Kennedy M, Enander RA, Tadiri SP, Wolfe RE, Shapiro NI, Marcantonio ER. Delirium risk prediction, healthcare use and mortality of elderly adults in the emergency department. J Am Geriatr Soc. 2014;62(3):462–469.

4. Ely EW, Gautam S, Margolin R, et al. The impact of delirium in the intensive care unit on hospital length of stay. Intensive Care Med. 2001;27(12):1892–1900.

5. Lewis LM, Miller DK, Morley JE, Nork MJ, Lasater LC. Unrecognized delirium in ED geriatric patients. Am J Emerg Med. 1995;13(2):142–145.

6. Trzepacz PT, Mittal D, Torres R, Kanary K, Norton J, Jimerson N. Validation of the Delirium Rating Scale-revised-98: comparison with the delirium rating scale and the cognitive test for delirium. J Neuropsychiatry Clin Neurosci. 2001;13(2):229–242.

7. Han JH, Wilson A, Graves AJ, et al. Validation of the Confusion Assessment Method for the Intensive Care Unit in older emergency department patients. Acad Emerg Med. 2014;21(2):180–187.

8. Han JH, Wilson A, Vasilevskis EE, et al. Diagnosing delirium in older emergency department patients: validity and reliability of the delirium triage screen and the brief confusion assessment method. Ann Emerg Med. 2013;62(5):457–465.

9. Luetz A, Weiss B, Boettcher S, Burmeister J, Wernecke KD, Spies C. Routine delirium monitoring is independently associated with a reduction of hospital mortality in critically ill surgical patients: A prospective, observational cohort study. J Crit Care. 2016;35:168–173.

10. Moon KJ, Lee SM. The effects of a tailored intensive care unit delirium prevention protocol: A randomized controlled trial. Int J Nurs Stud. 2015;52(9):1423–1432.

11. Pasin L, Landoni G, Nardelli P, et al. Dexmedetomidine reduces the risk of delirium, agitation and confusion in critically Ill patients: a meta-analysis of randomized controlled trials. J Cardiothorac Vasc Anesth. 2014;28(6):1459–1466.

12. Collins GS, Reitsma JB, Altman DG, Moons KGM, members of the Tg. Transparent Reporting of a Multivariable Prediction Model for Individual Prognosis or Diagnosis (TRIPOD): The TRIPOD Statement. Eur Urol. 2015;67(6):1142–1151.

13. Kersten H, Molden E, Willumsen T, Engedal K, Bruun Wyller T. Higher anticholinergic drug scale (ADS) scores are associated with peripheral but not cognitive markers of cholinergic blockade. Cross sectional data from 21 Norwegian nursing homes. Br J Clin Pharmacol. 2013;75(3):842–849.

14. Carnahan RM, Lund BC, Perry PJ, Culp KR, Pollock BG. The relationship of an anticholinergic rating scale with serum anticholinergic activity in elderly nursing home residents. Psychopharmacol Bull. 2002;36(4):14–19.

15. Sinoff G, Ore L. The Barthel activities of daily living index: self-reporting versus actual performance in the old-old (> or = 75 years). J Am Geriatr Soc. 1997;45(7):832–836.

16. Han JH, Zimmerman EE, Cutler N, et al. Delirium in older emergency department patients: recognition, risk factors, and psychomotor subtypes. Acad Emerg Med. 2009;16(3):193–200.

17. Jadhav A, Pramod D, Ramanathan K. Comparison of Performance of Data Imputation Methods for Numeric Dataset. Applied Artificial Intelligence. 2019;33(10):913–933.

18. Pedregosa F VG, Gramfort A et al. Scikit-learn: Machine Learning in Python. Journal of Machine Learning Research 12 (2011) 2825–2830.

19. Inouye SK, Westendorp RG, Saczynski JS. Delirium in elderly people. Lancet. 2014;383(9920):911–922.

20. Jorgensen SM, Carnahan RM, Weckmann MT. Validity of the Delirium Observation Screening Scale in Identifying Delirium in Home Hospice Patients. Am J Hosp Palliat Care. 2017;34(8):744–747.

21. Gavinski K, Carnahan R, Weckmann M. Validation of the delirium observation screening scale in a hospitalized older population. J Hosp Med. 2016;11(7):494–497.

22. Pendlebury ST, Lovett N, Smith SC, Cornish E, Mehta Z, Rothwell PM. Delirium risk stratification in consecutive unselected admissions to acute medicine: validation of externally derived risk scores. Age Ageing. 2016;45(1):60–65.

23. Lee S, Harland K, Mohr NM, et al. Evaluation of emergency department derived delirium prediction models using a hospital-wide cohort. J Psychosom Res. 2019;127:109850.

24. Wilson MG, Lee TC, Hass A, Tannenbaum C, McDonald EG. EMPOWERing Hospitalized Older Adults to Deprescribe Sedative Hypnotics: A Pilot Study. J Am Geriatr Soc. 2018;66(6):1186–1189.

25. Martin P, Tamblyn R, Benedetti A, Ahmed S, Tannenbaum C. Effect of a Pharmacist-Led Educational Intervention on Inappropriate Medication Prescriptions in Older Adults: The D-PRESCRIBE Randomized Clinical Trial. JAMA. 2018;320(18):1889–1898.

26. Wong BA YA, Liang AS, Gonzales R, Douglas VC, Hadley D. Development and Validation of an Electronic Health Record–Based Machine Learning Model to Estimate Delirium Risk in Newly Hospitalized Patients Without Known Cognitive Impairment. JAMA Network Open. 2018;1(4):e181018.

